# Exploring overcrowding trends in an inner city emergence department in the UK before and during COVID-19 epidemic

**DOI:** 10.1101/2021.01.20.21250150

**Authors:** J Panovska-Griffiths, J Ross, S Elkhodair, C Baxter-Derrington, C Laing, R Raine

**Affiliations:** Department of Applied Health Research, UCL, London, UK; Institute for Global Health, University College London, UK; The Queen’s College, Oxford University, Oxford, UK; Emergency Department, University College London NHS Foundation Trust, London, UK

**Keywords:** Emergency department, Healthcare quality improvement, Health services research, Statistics

## Abstract

**Background:** We compared impact of three pre-COVID-19 interventions and of the COVID-19 UK-epidemic and the first UK national lockdown on overcrowding within University College London Hospital Emergency Department (UCLH ED). The three interventions: target the influx of patients at ED (A), reduce the pressure on in-patients’ beds (B) and improve ED processes to improve the flow of patents out from ED (C).

**Methods:** We analysed the change in overcrowding metrics (daily attendances, the proportion of people leaving within four hours of arrival (four-hours target) and the reduction in overall waiting time) across three analysis. The first analysis used data 01/04/2017-31/12-2019 to calculate changes over a period of six months before and after the start of interventions A-C. The second and third analyses focused on evaluating the impact of the COVID-19 epidemic, comparing the first 10 months in 2020 and 2019, and of the first national lockdown (23/03/2020-31/05/2020).

**Results:** Pre-COVID-19 all interventions led to small reductions in waiting time (17%, p<0.001 for A and C;9%, p=0.322 for B) but also to a small decrease in the number of patients leaving within four hours of arrival (6.6%,7.4%,6.2% respectively A-C,p<0.001).

In presence of the COVID-19 pandemic, attendance and waiting time were reduced (40% and 8%;p<0.001), and the number of people leaving within four hours of arrival was increased (6%,p<0.001). During the first lockdown, there was 65% reduction in attendance, 22% reduction in waiting time and 8% increase in number of people leaving within 4 hours of arrival (p<0.001). Crucially, when the lockdown was lifted, there was an increase (6.5%,p<0.001) in the percentage of people leaving within four hours, together with a larger (12.5%,p<0.001) decrease in waiting time. This occurred despite the increase of 49.6%(p<0.001) in attendance after lockdown ended.

**Conclusions:** The mixed results pre-COVID-19 (significant improvements in waiting time with some interventions but not improvement in the four-hours target), may be due to a ‘spill-over effect’ where clogging up one part of the ED system affects other parts. Hence multifaceted interventions and a system-wide approach to improve the pathway of flow through the ED system is necessary.

During 2020 and in presence of the COVID-19 epidemic, a shift in public behaviour with anxiety over attending hospitals and higher use of virtual consultations, led to notable drop in UCLH ED attendance and consequential curbing of overcrowding.

Importantly, once the lockdown was lifted, although there was an increase in arrivals at UCLH ED, overcrowding metrics were reduced. Thus, the combination of shifted public behaviour and the restructuring changes during COVID-19 epidemic, maybe be able to curb future ED overcrowding, but longer timeframe analysis is required to confirm this.

## Introduction

The spread of COVID-19 during 2020 has put pressure and induced a number of changes to the National Health Service (NHS) in the UK. To prevent overwhelming the health services, including Emergence Departments (EDs) and to suppress the increasing number of COVID-19 cases in the UK earlier this year, the UK Government imposed strict social distancing measures (“lockdown”) from March 23, 2020. As the number of cases started to decline following the lockdown and over the following months, a phased relaxing of the lockdown measures started with primary schools opening on June 01, 2020 and wider relaxing of imposed measures from July 4, 2020. After remaining low during July and August, the number of COVID-19 cases and associated deaths, started to increase again in late August and throughout September 2020. After trialling a Tier system for more localised strict control measures, which was insufficient to curb the large recent resurgence in cases and COVID-19 associated deaths, a second national lockdown was imposed across the UK from November 5, 2020.

EDs provide immediate assessment and care to patients, who may be critically ill. During the first wave of COVID-19, the level of visit to EDs declined sharply; 48% reduction was reported nationally in April 2020 compared to April 2019 [47]. While awaiting roll-out of an effective vaccine, maintaining social distancing, in conjunction with effective testing, tracing and isolation of positive cases, are the main interventions for virus suppression. Maintaining social distancing is difficult to achieve in overcrowded EDs.

ED treatment in the pre COVID-19 era was often hindered by overcrowding. Contributing factors include seasonal increases in demand, onset of an epidemic or pandemic [6,7], shortage of available inpatient beds, staff shortage [8,9] and inefficient patient flow through the system [10,11]. Previous studies have shown that overcrowding in ED is associated with increased mortality, increased length of stay, reduced quality of care, poor patient experience and increased number of serious incidents [1-4]. However interventions have had varying levels of success [5], and despite ongoing efforts, there is understanding on how to improve ED patient flow [4].

The Royal College of Emergency Medicine suggests that interventions focusing on changes in the input, throughput and output parts of the ED system may be one way of relieving overcrowding [17-19]. Input interventions involve targeting aspects responsible for managing the number of patients attending the ED e.g. relocation of primary care services within emergency care services [20]. However the evidence for this is weak, limited and outdated [21]. Throughput interventions comprise processes within the ED and can include aspects such as staffing, co-ordination with inpatient teams, local protocols and the physical layout of the department. For example training nurses to order an X-rays at triage [22] is a throughput intervention. Finally, output interventions focus on improving the exit of patients from the emergency department, through either admission, discharge or transfer to another service. For example, boarding patients, i.e. sending them to an inpatient ward while waiting for a bed is an output intervention [23,24]. However, the evidence base of the effectiveness of these interventions is small, limited and heterogeneous [25,26].

To improve emergency care provision and reduce prolonged inpatient length of stay, the UK Department of Health and Social Care in 2004 produced a white paper [12] that set a standard target for acute hospitals as part of the National Health Service (NHS) that at least 95% of patients attending EDs must be seen, treated, admitted or discharged in under four□hours. This still represents the UK’s key indicator of an ED performance [13].

Modelling can aid assessment of the impact of different interventions. To date a number of studies have used modelling to quantify the impact of ED system changes on ED overcrowding as outlined in recent reviews for the UK system [27] or the USA system [28]. Different modelling approaches such as statistical (regression) modelling, mathematical (i.e. queueing-theory based) modelling and discrete event simulations can be used for this purpose and have different advantages and disadvantages. Generally, statistical modelling is used to evaluate impact of implemented interventions, queueing theory can evaluate flow of people through the ED system and hence explore impact of potential new interventions, while the discrete event simulators allow tracking of individual patents through the system and hence identify potential “clogs” within the system. In existing studies, specific type of models are constructed for the posed question, but often the details of the modelling framework have not always been transparent and difficult to replicate.

In this paper we examine overcrowding in the ED within the University College London Hospitals NHS Foundation Trust, UCLH. Our study is using data from this ED over a long period before the COVID-19 epidemic (April 01, 2017-December 31, 2019) as well as during the first ten months of the COVID-19 epidemic in 2020. We compare the impact on overcrowding of three trialled interventions at different times pre-COVID-19 to the impact of the presence of the epidemic and the imposed first national lockdown to suppress it and protect the NHS. The three interventions targeted the influx of patients at ED (interventions A), reduced the pressure on in-patients’ beds (interventions B) or improved internal ED processes to increase outflow of patents from ED (interventions C) (details in Methods).

## METHODS

### Setting

The study was conducted in the Emergency Department of the University College London Hospital (UCLH ED) over the study period April 01, 2017 - October 14, 2020. All patients aged 16 and above attending UCLH ED over this period were included. The study protocol was performed in accordance with the relevant guidelines.

### Interventions

Since April 2017, three groups of interventions targeting different aspects of the UCLH ED have been implemented in timeline shown in Figure 1. The interventions and their evidence base are summarised in Table 1.

**Table 1:** Description and evidence base for different interventions A-C implemented at UCLH ED over the study period with timeline given in Figure 1.

| Intervention target | Intervention name (start date) | Type of intervention | Summary on extend/strength of intervention |
| --- | --- | --- | --- |
| Targeting the influx of patients | Interventions A:<br>New triage setting with co-located GP and presence of senior doctor at UCLH ED (February 2018) | Input stream | There is some evidence that changes to triage such as co-locating GPs within triage, having a senior doctor at triage or having rapid assessment pods within triage, can be effective in redirecting the flux of incoming patients and lead to reduced ED waiting time [20,21,27,29-33]. |
| Reducing pressure upon available patients' bed | Interventions B (October 2017)<br>Stream B1: Expansion of emergency floor<br>Stream B2: : Same day emergency clinic<br>Stream B3: Medical EAU | Throughput stream<br>Output stream<br>Output stream | There is some evidence that increasing bed numbers within ED can reduce ED waiting time [34-36].<br>There is some evidence that having the option to redirect patients arriving at triage to primary care, via same day emergency clinic may reduce overall ED waiting time [4,27].<br>There is some evidence that presence of acute medical units within EDs can reduce ED waiting times [37-39]. |
| Improving internal processes to increase outflow | Interventions C: (April 2019)<br>Facilitation the workup of patients to include three streams: optimisation of workforce, clinical pathway and full digitisation of ED | Throughput streams | There is some evidence that interventions including improving specialty in-reach to ED or GPs working within the ED [4,20,33] or boarding patients [42] or digitization of the records within the Electronic Health Records (EHRs) database [41] can reduce ED overcrowding. |

**Figure 1:**
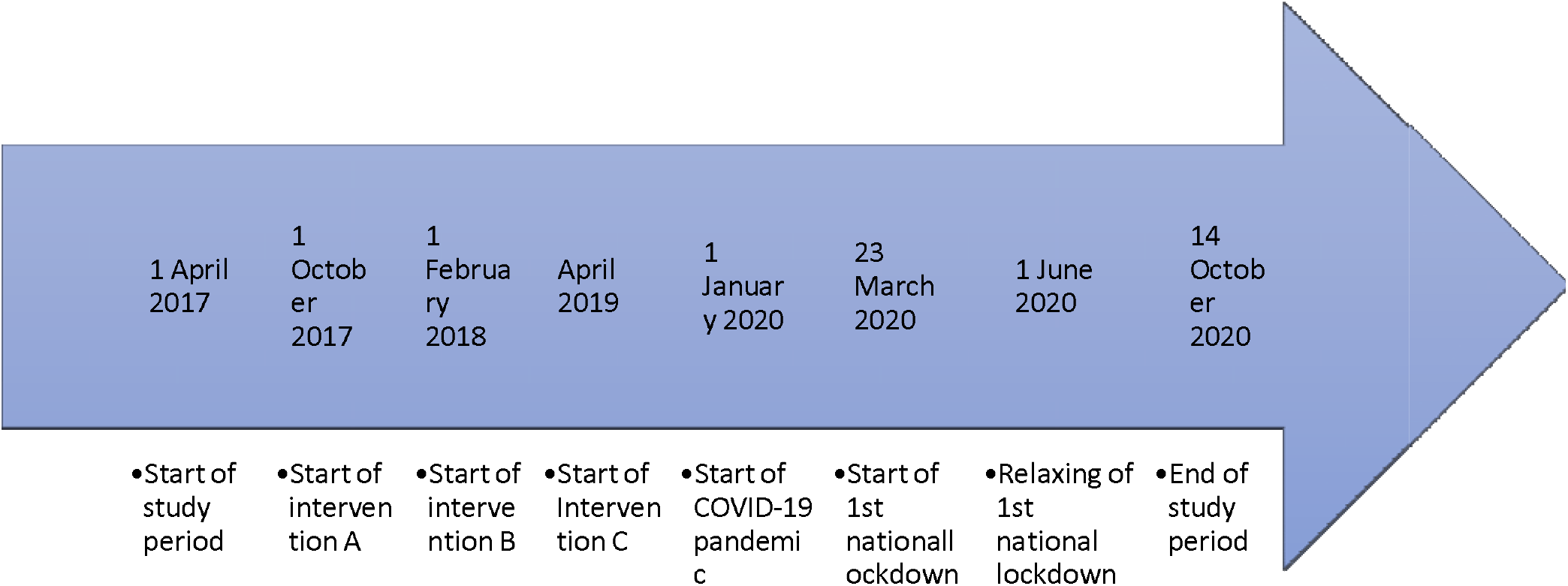
Timeline of different interventions within the study period

Interventions A are input interventions that target the pathways of walk-in patients by implementing a triage system where walk-in patients are signposted and diverted directly to other services, including redirecting to community care and directly referred to medical/surgical assessment units. These interventions began in February 2018.

Interventions B aim to reduce the pressure upon available in-patients’ beds. They constitute three streams: one throughput stream (B1) that focuses on expansion of the emergency department by adding more mobile beds, and two output streams (B2 and B3) that introduce the same day emergency clinic and a focus on medical emergency assessment unit (EAU). Stream B2 focuses on an expansion of the ambulatory emergency care unit to assess ambulant patients being considered for emergency admission. Stream B3 encompasses opening of an EAU used by the acute medical team as a place to clerk and admit ED patients, in order to free up beds in ED and streamline the medical admission process. It is co-located with the ambulatory emergency care unit, allowing patients to be transferred between them as appropriate. These interventions were established in October 2017.

Finally, interventions C represent throughput interventions that aim to improve ED processes and hence increase flow of patents out from ED. They comprise three streams: optimisation of the workforce by for example having senior or speciality doctors with ED in-reach or having co-located GPs within ED, having same-day emergency clinics and introduction of a digitisation of health records. This revolves around the use of the Electronic Health Records (EHRs) database as a fully electronic health record system that has been introduced across the Trust including the UCLH ED. This allows single sign on for clinicians and a consistent health record for patients replacing paper notes and prescription charts. Interventions C began in April 2019.

### Design

Interrupted time series analysis (ITS) and regression analysis.

### Data

Routine hourly data on the number of patients arriving at UCLH ED over the study period of 42 months were collated. This period includes six months (April 01, 2017-September 30, 2017) before any interventions were implemented and a period of eight months after the last intervention was implemented (April, 01 2019-December 31, 2020). It also includes the first 10 months of the UK COVID-19 epidemic in 2020 (January 01, 2020-October 14, 2020). People arriving at UCLH ED were grouped into people walking into UCLH ED or arriving by ambulance.

### Statistical analysis

We constructed time series of the daily arrivals of patients at UCLH ED over the period April 01, 2017-October 14, 2020. Using the data, we truncated the observation period into two periods: April 01, 2017-December 31, 2020 and January 01,2020-October 14,2020; the formed period representing pre-COVID-19 era and the latter COVID-19 period in the UK in 2020 for which we have data. The pre-COVID-19 era included periods of before and after intervention for each of the three sets of interventions A-C. The COVID-19 period included the period in 2020 during the first national lockdown (March 23, 2020-May 31, 2020) (‘lockdown’) and the period before and after the first COVID-19 epidemic wave. The timeline of our work is schematically represented in Figure 1. The dataset gave us sufficient power for statistical analysis.

Using these different time periods of the available data we undertook three separate but linked analysis.

The first analysis focused on the impact of trialled interventions in the pre-COVID-19 era on overcrowding at UCLH ED. For this analysis we used the timeseries between April, 01, 2017 and December 31, 2020 (inclusive). Over this period we defined the start of each set of interventions A, B and C; respectively February 01, 2018, October 01, 2017 and April 01, 2019. We then calculated the changes in the overcrowding metrics over a period of six months before and after the start of each intervention.

The second and third analysis focused on evaluating the impact of the COVID-19 epidemic and the first national lockdown on overcrowding in UCLH ED.

The second analysis compared the number of attendances, % of people leaving within 4 hours of arrival and the average daily waiting time over the first 10 months of 2019 and 2020 (January 01-October 14) i.e. in absence and presence of the COVID-19 epidemic. Additionally we also compared the first national lockdown period (March 23, 2020-May 31, 2020) in both years. By looking at all three overcrowding metrics, this analysis informed whether overcrowding changes were a result of lowered attendances at this ED during 2020 or a consequence of COVID-19-induced changes here. We note that to accommodate COVID-19 safe environment, a number of changes were made in this ED during the first epidemic wave e.g. rapid assessment and triage were stopped and adaptations were made to facilitate more rapid flow of patients through the ED (e.g. segregated pathways, streaming patients, some patients went in escalated treatments such as non-invasive ventilation before leaving the ED etc).

For the third analysis, we split the data from 2020 into three time periods of 71 days defined as period before the first national lockdown (January 12, 2020-March 22, 2020) (‘before lockdown’), period during the first national lockdown (March 23, 2020-May 31, 2020) (‘lockdown’) and period after the first national lockdown (June 01, 2020-August 11, 2020)(‘after lockdown’). For each of these periods we compared the number of attendances, % of people leaving within four hours of arrival and the average daily waiting time. This analysis informed whether lockdown significantly affected overcrowding in this ED.

For each analysis, we used mixed effects negative binomial regression models to quantify the changes in the three overcrowding metrics, projecting the incidence rate ratios (IRRs) for all people attending UCLH ED as well as split between those arriving by ambulance and walking in, the % of people leaving within four hours and the average time spend in UCLH ED (henceforth waiting time). To account for differences associated with the intervention period, we fitted a random intercept model with a single indicator variable for “during-intervention/lockdown” for each intervention and the lockdown. To estimated standard errors we used bootstrapping with 500 replications and also reported the 95% confidence intervals (95% CI) and p-values. All the statistical analysis was undertaken in Stata v16.

## RESULTS

### Descriptive statistics over the study period

Figures 2(a) and 2(b) contain, respectively, the timeseries of the attendances at UCLH ED and the percentage of people that left within four hours of arrival in the period between April 1, 2017 and October 14, 2020. We observe that there was almost constant level of attendance at this ED in the period before March 2020 with a notable drop in attendances that coincides with the onset of the first national lockdown (23/03/2020).

**Figure 2(a)-(b):**
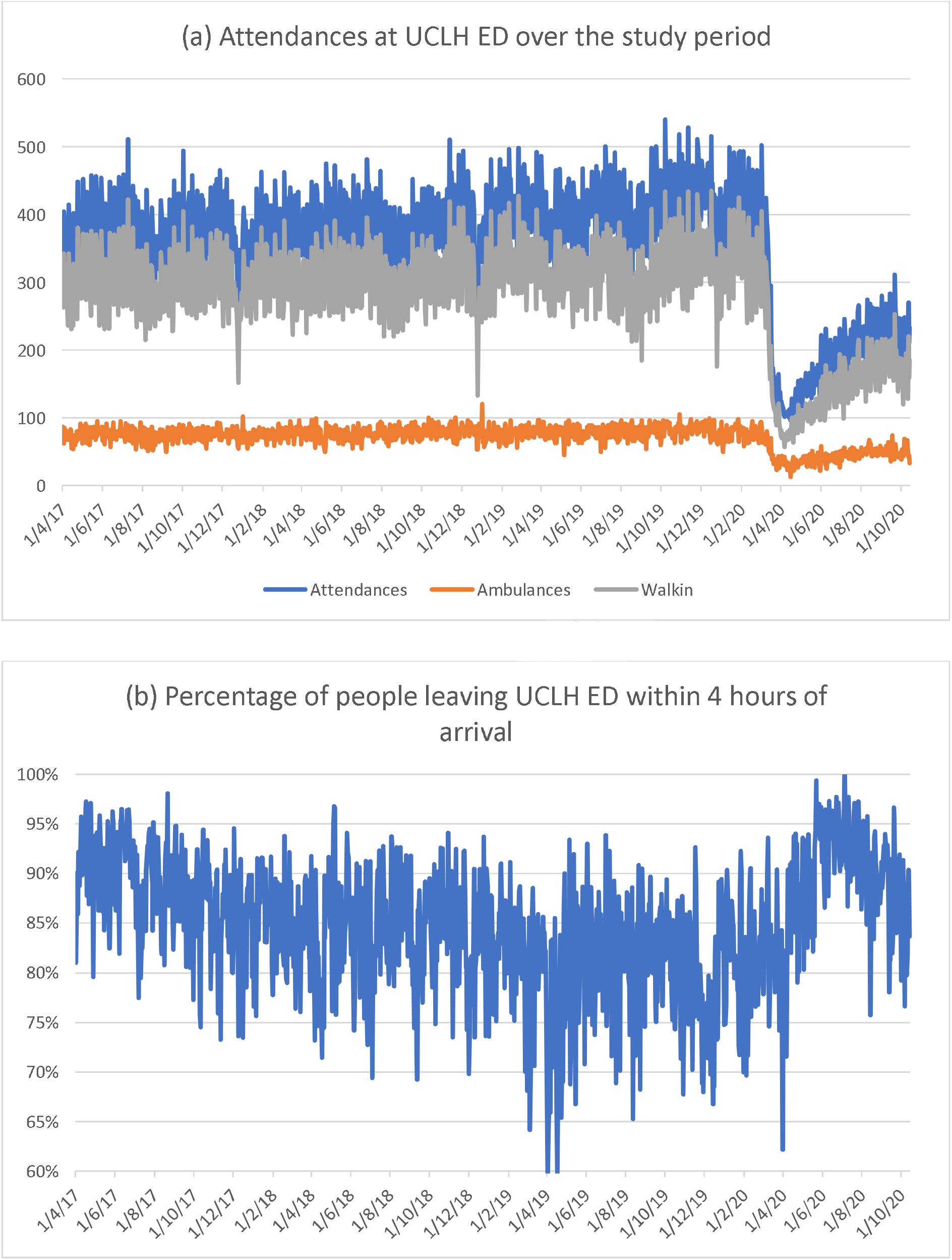
(a) Timeseries of number of people attending UCLH ED over the study period stratified by people arriving by ambulance and walking-in. There is a large drop in attendances that coincides with the imposing of the first national COVID-19 lockdown. While this drop is evident for both walk-in and ambulance patients, it is larger in walk-in patients. (b) Timeseries of the number of people leaving UCLH ED within 4 hours of arrival.

Over the entire study period, a total of 458,456 patients attended UCLH ED of whom 92,362 (20.1%) arrived by ambulance and 366,094 (79.8%) patients walked-in. Daily, on average 355 patients attended UCLH ED, with 72 patients arriving by ambulance and an average of 283 patients walking-in. The average time spend at UCLH ED was 3.5 hours (95%CI=[3.3,3.6]), with times spend by patients coming by ambulance spending on average 4.6 hours (95%CI=[4.3,4.9]), while patients walking-in spending, on average, 3.2 hours (95%CI=[3.0,3.4]) at UCLH ED. Finally, the average proportion of people leaving UCLH ED within 4 hours of arrival was 84.8% (95%CI=[84.4%,85.2%]).

### Impact of trialed interventions in the pre-COVID-19 era

The results of the first analysis showing the impact of the overcrowding interventions A-C on the overcrowding metrics are contained in Table 2.

**Table 2:**
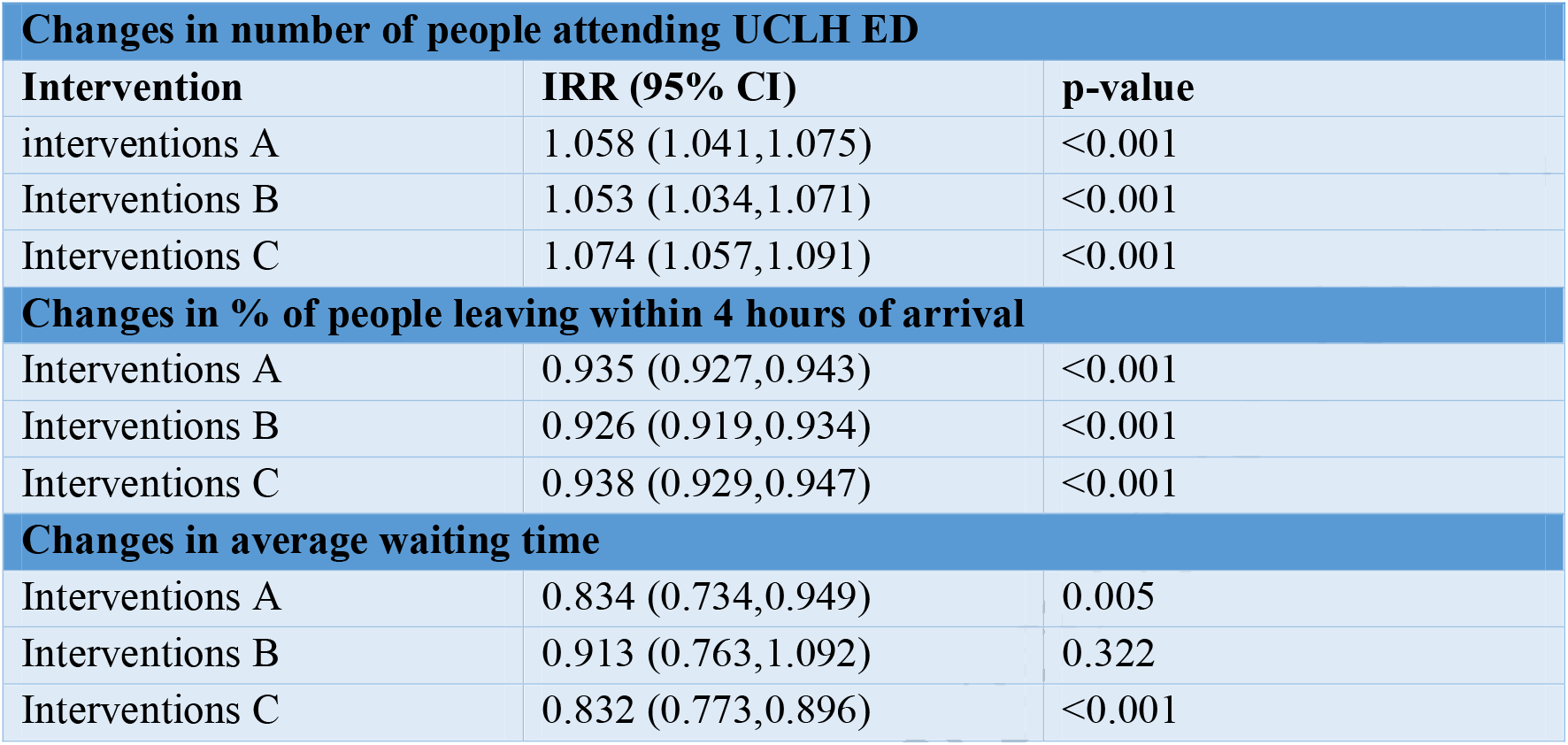
Outcomes from the ITS analysis projecting the impact of interventions A, B and C on the overcrowding indicators. The study period is truncated into periods before and after the start of each intervention A, B and C; respectively starting on February 01, 2018, October 01, 2017 and April 01, 2019. We calculate the changes over a period of six months before and after the start of each intervention.

| **Changes in number of people attending UCLH ED** |  |  |
| --- | --- | --- |
| **Intervention** | **IRR (95% CI)** | **p-value** |
| interventions A | 1.058 (1.041,1.075) | <0.001 |
| Interventions B | 1.053 (1.034,1.071) | <0.001 |
| Interventions C | 1.074 (1.057,1.091) | <0.001 |
| **Changes in % of people leaving within 4 hours of arrival** |  |  |
| Interventions A | 0.935 (0.927,0.943) | <0.001 |
| Interventions B | 0.926 (0.919,0.934) | <0.001 |
| Interventions C | 0.938 (0.929,0.947) | <0.001 |
| **Changes in average waiting time** |  |  |
| Interventions A | 0.834 (0.734,0.949) | 0.005 |
| Interventions B | 0.913 (0.763,1.092) | 0.322 |
| Interventions C | 0.832 (0.773,0.896) | <0.001 |

The number of people attending UCLH ED increased slightly, albeit statistically significant, in the period after implementation of interventions A, B and C (table 2, rows 3-5). The increase in numbers of patients attending UCLH ED after implementation of interventions A (row 3 of Table 2) was 5.8%, for interventions B was 5.3% (row 4 of Table 2) and for interventions C was 7.4% (row 5 of table 2).

There was a small but statistically significant decrease in number of people leaving within four hours of arrival (rows 7-9 of Table 2). For interventions A this reduction was 6.6% (p<0.001), from 88.5% to 82%. For interventions B this reduction was 7.4% (p<0.001) from 90.4% to 83% of people leaving within 4 hours of arrival (Table 2). Finally, for interventions C this reduction was 6.2% (p<0.001), from 86.2% to 80% of people leaving within four hours of arrival.

The average waiting time spend by patients at UCLH ED was reduced by 38 minutes (17%) after implementation of interventions A (p=0.005) and C (p<0.001) (rows 11 and 13 of Table 2 respectively). With implementation of intervention B, the average waiting time was reduced by 20 minutes (9%) but this was not statistically significant (p=0.322 in row 12 of Table 2).

### Impact of COVID-19 on overcrowding

The results of the second and third analysis showing the impact of the epidemic and the lockdown on overcrowding metrics in this ED are shown in Table 3.

**Table 3:**
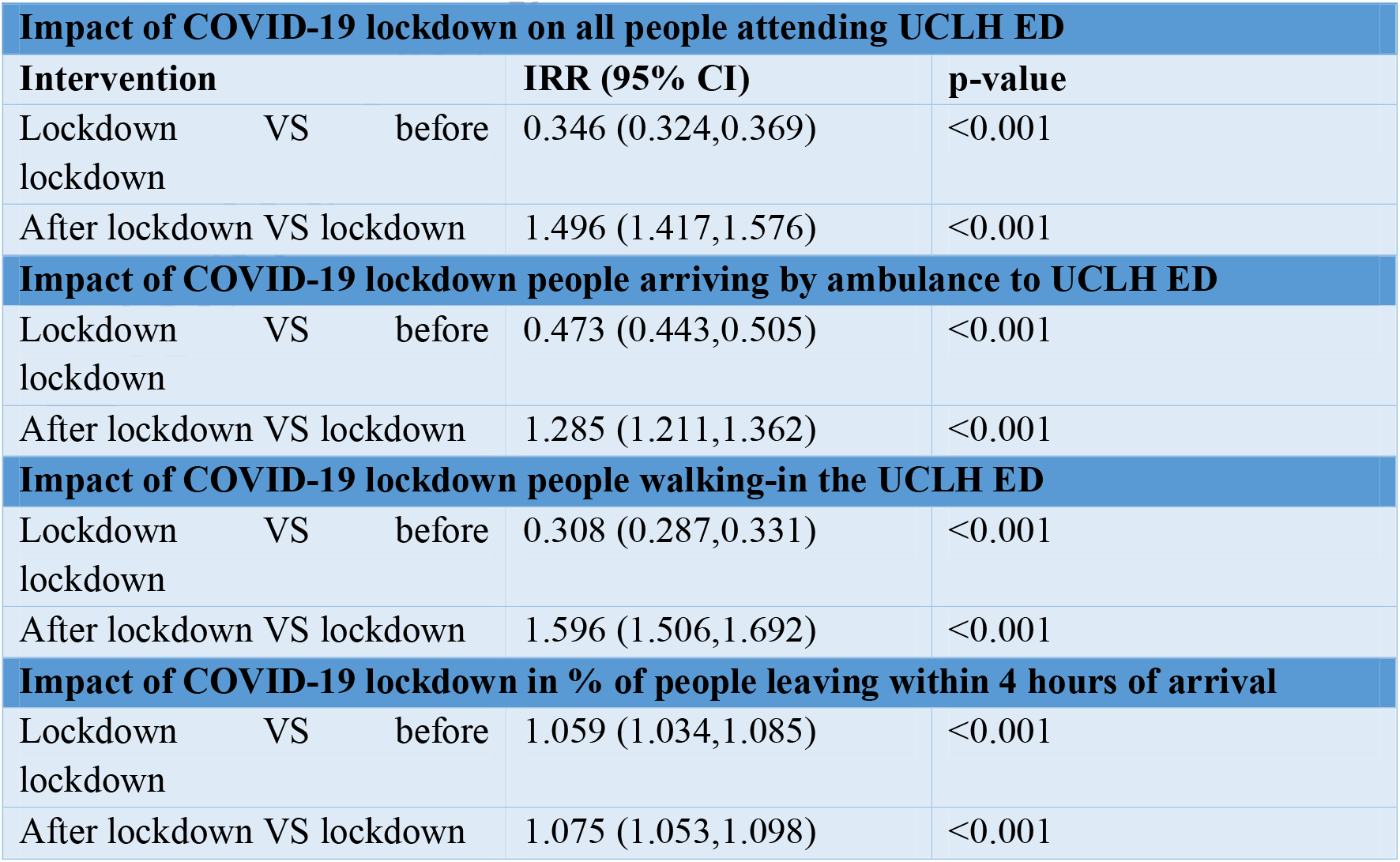
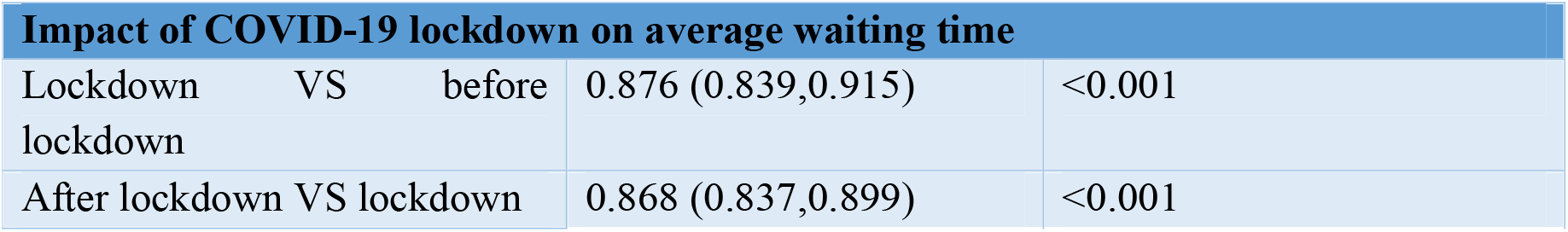
Outcomes from the ITS analysis projecting the impact of the first national lockdown (‘lockdown’) to suppress COVID-19 during the spring of 2020 on the overcrowding metrics and considering all attendances and those arriving by ambulance or walking in. The study period (over the period January 12, 2020-August 11, 2020) is split into three time periods of 71 days defined as before the first national lockdown (January 12, 2020-March 22, 2020) (‘before’), during the first national lockdown (March 23, 2020-May 31, 2020) (‘lockdown’) and after the first national lockdown (June 01, 2020-August 11, 2020)(‘after’).

| **Impact of COVID-19 lockdown on all people attending UCLH ED** |  |  |  |  |
| --- | --- | --- | --- | --- |
| **Intervention** |  |  | **IRR (95% CI)** | **p-value** |
| Lockdown VS before lockdown |  |  | 0.346 (0.324,0.369) | <0.001 |
| After lockdown VS lockdown |  |  | 1.496 (1.417,1.576) | <0.001 |
| **Impact of COVID-19 lockdown people arriving by ambulance to UCLH ED** |  |  |  |  |
| Lockdown VS before lockdown |  |  | 0.473 (0.443,0.505) | <0.001 |
| After lockdown VS lockdown |  |  | 1.285 (1.211,1.362) | <0.001 |
| **Impact of COVID-19 lockdown people walking-in the UCLH ED** |  |  |  |  |
| Lockdown VS before lockdown |  |  | 0.308 (0.287,0.331) | <0.001 |
| After lockdown VS lockdown |  |  | 1.596 (1.506,1.692) | <0.001 |
| **Impact of COVID-19 lockdown in % of people leaving within 4 hours of arrival** |  |  |  |  |
| Lockdown VS before lockdown |  |  | 1.059 (1.034,1.085) | <0.001 |
| After lockdown VS lockdown |  |  | 1.075 (1.053,1.098) | <0.001 |

Table 3: Outcomes from the ITS analysis projecting the impact of the first national lockdown ('lockdown') to suppress COVID-19 during the spring of 2020 on the overcrowding metrics and considering all attendances and those arriving by ambulance or walking in.
| Impact of COVID-19 lockdown on average waiting time |  |  |  |  |
| --- | --- | --- | --- | --- |
| Lockdown VS before lockdown |  |  | 0.876 (0.839,0.915) | <0.001 |
| After lockdown VS lockdown |  |  | 0.868 (0.837,0.899) | <0.001 |

Our second analysis suggested that in the presence of the COVID-19 epidemic, overcrowding in UCLH ED was reduced. During 2020, there was a notable drop in the number of attendances at UCLH ED that coincides with the onset of the first national lockdown (Figure 2(a) and Table 3). While drop in attendances was evident in both people walking in UCLH ED and people arriving with ambulances, the decrease in those walking in was larger (Figure 2(a) and Table 3)

Specifically, during the period January 01, 2020-October 14, 2020, a total of 69,491 patients attended UCLH ED of which 14,837 (21.4%) arrived by ambulance and 54,654 (78.63%) patients walked-in. This was a 39.3% reduction in visits to this ED in comparison to the same period in 2019. In addition, comparing these periods in 2019 and 2020, we determined that there was 8% reduction in waiting time and 6% increase in number of people leaving within four hours of arrival (Table 3).

Considering only the period of the first national lockdown (March 23-June 01) in 2019 and 2020, these changes were even more dramatic: 65.3% reduction in attendance and 21.9% reduction in waiting time and 8.3% increase in number of people leaving within four hours of arrival (Table 3).

For the third analysis, considering 2020 only, our results suggest that the lockdown significantly affected the three overcrowding metrics. In comparison to the period before the lockdown, the lockdown led to 65.4% reduction in attendance (52.7% in those arriving by ambulance and 69.2% in those walking in), 12.4% reduction in waiting time and 5.9% increase in number of people leaving within four hours of arrival; all statistically significant (Table 3).

Once the strict social distancing measures were lifted, there was an 49.6% increase in attendances to UCLH ED (28.5% in those arriving by ambulance and 59.6% in those walking in) (Table 3). But interestingly, compared to during lockdown, in the period afterwards, the waiting time was reduced by 13.2%, while the number of people leaving within four hours of arrival was increased by 7.5%; all changes statistically significant (Table 3).

## DISCUSSION

All three sets of interventions A-C trialled in the pre-COVID-19 era led to small reductions of waiting time over the two years before the epidemic. These reductions were significant in the case of interventions A (that targeted the influx of patients at ED) and interventions C (that improved internal ED processes to increase outflow of patents from ED) but were not in the case of interventions B (that aimed to reduce the pressure on in-patients’ beds). In addition, all three trialled interventions were also associated with a small, but significant decrease in the number of patients leaving the ED within four hours of arrival. Therefore with the trialled interventions pre-COVID-19 no improvement was evident in the four-hours target.

During 2020 and in presence of the COVID-19 pandemic, there was a drop in attendance at this ED and as a result, waiting times were reduced and the percentage of people leaving within four hours or arrival increased. This result is unsurprising: the drop in attendances at UCLH ED compared to the same periods in 2019 reflects the national data [47,48]. Crucially, however when the lockdown was lifted following the first wave of COVID-19, there was both an increase (6.5%) in the percentage of people leaving within four hours, together with a larger (12.5%) decrease in waiting time. This occurred despite the increase of 49.6% in ED attendance after lockdown ended.

The mixed results found pre-COVID-19 (significant improvements in waiting time with some interventions but fewer people leaving UCLH ED within four hours of arrival), may be due to a ‘spill-over effect’. For example, clogging up one part of the ED system e.g. triage will have a knock on effect and increased pressure on the outflow of patients. This is simply due to the fact that large influx will produce a bottle neck effect within the system. In a situation where there is limited capacity in terms or inpatients’ beds or a complex system of discharging from hospitals, this will lead to lower outflow through the system i.e. elevated overcrowding. Therefore, in line with existing literature [43,44], there is a need for the entire pathway of flow through the ED system to be examined and a system-wide improvement to be implemented. This needs to include separate parts (a) deciding the priority measure of overcrowding (b) identifying the stream of blockages within different parts that affect it and taking an informed approach with engagement of staff to design and implement a modular strategy.

Our findings reflect those in the literature. For example, a systematic review [42] of the effectiveness of interventions on reducing ED crowding by older patients between 1990 and 2017 showed that none of the implemented interventions reduced all overcrowding measures. Our findings of the need for multifaceted interventions and a system-wide approach is also supported by the literature [4]. For example, results in [42] suggest that rapid assessment and streaming of care for older adults arriving at the ED lead to a statistically significant decrease of ED length of stay. This is analogous to how implementation of interventions A led to statistically significant reduction of the time spend in ED in our study. Similarly, existing work showed that the presence of an ED-based consultant geriatrician was associated with significant time reduction between patient admission and geriatric review compared to an in-reaching geriatrician [43]. This has parallels with our findings that implementation of interventions C leads to reduction to the time spend in ED.

Our results also reflect the existing literature on the impact of COVID-19 on ED overcrowding [47, 49-50] and add to it. For example, our findings of the drop in visit to EDs during the first lockdown, reflect the nationally reported decline of 48% reduction in attendances in April 2020 compared to April 2019 [47,48]. The drop in attendance during the first stages of the pandemic spread, compared to these being stable in the pre-COVID-19 period, are also in agreement with the findings from a study in the inner city ED of the University Hospital of Parma, Italy [49] and across 24 EDs from five American states [50]. The literature across these studies agrees that the COVID-19 pandemic lead to reduced attendance of EDs and as a consequence overcrowding was curbed.

A shift in public behaviour during the first wave of the pandemic can explain these results. For example, during the first lockdown period in the UK, when the public was instructed to ‘stay at home, save lives and protect the NHS’, anxiety over presenting in hospitals and higher use of NHS 111 and other advice lines occurred [47]. Nationally, this led to a decline in number of arrivals within EDs with minor injuries [47,48]. This allowed ambulance services to more effectively respond to people with more severe needs. In addition, primary care services made a substantive shift to virtual primary care consultations [48], which gave immediate access to patients with less severe injuries. Hence, the reduction in overcrowding in 2020 compared to 2019, would have been due to changes in the ways in which people with less severe needs gained immediate access to services virtually and hence attended EDs less frequently, leading to more appropriate use of the ED services.

After the first COVID-19 wave, an increase was evident in attendances in this ED and notably from patients walking in (Figure 2(a)). Despite this, waiting time at UCLH ED was reduced while the number of people leaving UCLH ED within four hours of arrival was increased. We note two things. Firstly, the increase in UCLH ED attendance after the first lockdown has not yet reached baseline pre-COVID-19 level (Figure 2(a)). Secondly, a large restructuring of EDs, including UCLH ED has occurred as part of the COVID-19 epidemic response. Hence, the combination of shifted public behaviour of more appropriate use of EDs and the system-wide restructuring changes during COVID-19 epidemic would have increased flow of patients in this ED and hence curb ED overcrowding during 2020. It is important to assess whether this remains the case in future, and while our study is the first to study thus, we are aware that a longer timeframe analysis is necessary to confirm this.

The choice of statistical modelling in this study has some limitations. Our method, interrupted time series, can effectively compare changes in outcomes for successive groups of patients before and after ED interventions/lockdown started, and is therefore fit for the purpose of this study. But whilst such regression modelling is useful in drawing conclusion for the duration of the study where fitted curves mimic the data, the presence of turning points in the non-linear fits makes them unreliable for prediction beyond the period for which data are available. An alternative would be to develop and utilise queuing theory models e.g.[45], that model the flow of patients through a system.

Another possible future direction in research would be use of automated learning systems, rooted in higher-order statistical methods e.g. machine learning, that will allow real-time assessment of patients arriving at ED. A recent review [46] has collated the existing literature on the application of AI methods in emergency medicine, concluding that although some attempts have been made in the field, this an emerging field that will benefit from application of tailor-made machine learning algorithms for real-life assessment of, for example, arrival and triage assessment. Future work could combine the use of Electronic Health Records and robust machine learning algorithms.

## CONCLUSIONS

In summary, our study found that trialled interventions pre-COVID-19 lead to mixed results: significant improvements in waiting time with interventions that targeted the influx of patients at ED (A) or improved internal ED processes to increase outflow of patents from ED (C), but fewer people leaving UCLH ED within four hours of arrival. A spill-over effect where clogging up one part of the ED system may have an effect on other parts and overall, would have been responsible for this. Hence, there is a need for multifaceted interventions and a system-wide approach to improve the entire pathway of flow through the ED system.

During 2020 and in presence of the COVID-19 epidemic, a shift in public behaviour with anxiety over presenting in hospitals and higher use of NHS 111 or virtual primary consultations, UCLH ED attendance dropped notably, and as a consequence waiting time was reduced while the percentage of people leaving the ED within four hours was increased.

Importantly, once the lockdown was lifted, there was an increase in arrivals at UCLH ED, but not yet reaching the pre-COVID-19 attendance level. As a result, even with this increase the overcrowding metrics were reduced. Therefore, it may be possible that shift in public behaviour to more appropriately use EDs combined with the system-wide restructuring changes during COVID-19 epidemic would be able to curb overcrowding in future, but longer timeframe analysis is required to attest this.

## Data Availability

The datasets used and analysed during this study and the numerical codes used to generate the outcomes of this paper are available from the corresponding author on reasonable request.

## Abbreviations

Not applicable

## Declarations

### Ethics approval and consent to participate

This study is considered a retrospective service evaluation and as such is exempt and ethics approval and informed consent for current study is waived by the UCL Research Ethics Committee.

### Consent for publication

Not applicable.

### Authors’ contributions

JPG had the idea for the study, undertook the statistical analysis and drafted the manuscript. Data was supplied by CBD. Collation of existing interventions was done by JPG, JR and CE with input from CL. The manuscript was written by JPG, with contributions from JR, RR, SE CBD and CL to manuscript’s revision and refinement. All authors read and approved the final manuscript.

## Acknowledgements

Not applicable

